# Early trends for SARS-CoV-2 infection in central and north Texas and impact on other circulating respiratory viruses

**DOI:** 10.1101/2020.04.30.20086116

**Authors:** Manohar B. Mutnal, Alejandro C. Arroliga, Kimberly Walker, Amin Mohammad, Matthew M. Brigmon, Ryan M. Beaver, John K. Midturi, Arundhati Rao

## Abstract

**Introduction:** Rapid diagnosis and isolation are key to containing the quick spread of a pandemic agent like *SARS-CoV-2*, which has spread globally since its initial outbreak in Wuhan province in China. *SARS-CoV-2* is novel to most parts of the world including USA and the effect on typically prevalent seasonal viruses is just becoming apparent. We present our initial data on the prevalence of respiratory viruses in the month of March, 2020.

**Methods:** This is a retrospective cohort study post launching of *SARS-CoV-2* testing at Baylor Scott and White Hospital (BSWH), Temple TX. Testing for *SARS-CoV-2* was performed by real-time rRT-PCR assay and results were shared with State public health officials for immediate interventions.

**Results:** More than 3500 tests were performed during the first two weeks of testing for *SARS-CoV-2* and identified 168 (4.7%) positive patients. Sixty-two (3.2%) of the 1,912 ambulatory patients and 106 (6.3%) of the 1,659 ED/inpatients were tested positive. Higher rate of infection (6.9%) were noted in the patients belonging to age group 25-34 years and least number of positive cases were noted in <25 years old (2%) group. The TX State county specific patient demographic information was shared with respective public health departments for epidemiological interventions.

Incidentally, this study showed that there was a significant decrease in the occurrence of infections due to seasonal respiratory viruses in this region, perhaps due to increased epidemiological awareness, about *SARS-CoV-2*, among general public. Data extracted for BSWH from the CDC’s National Respiratory and Enteric Virus Surveillance System (NREVSS) site revealed that Influenza incidence declined to 8.7% in March 2020 compared to 25% in March 2019.

**Conclusions:** This study was intended to provide an initial experience of dealing with a pandemic and the role of laboratories in crisis management. This study provided *SARS-CoV-2* testing data from ambulatory and inpatient population. Epidemiological interventions depend on timely availability of accurate diagnostic tests and throughput capacity of such systems during large outbreaks like *SARS-CoV-2*.

## Introduction

In December 2019, Wuhan city, the capital of Hubei province in China, became the center of an outbreak of pneumonia of unknown cause. By Jan 7, 2020, Chinese scientists had isolated a novel coronavirus, severe acute respiratory syndrome coronavirus 2 (*SARS-CoV-2*; previously known as 2019-nCoV), from these patients with virus-infected pneumonia.^1^ Cases have now spread to 190 countries. As of March 23, 2020 there were more than 372,000+ confirmed cases and 16,000+ deaths.^2^ Although the outbreak is likely to have started from a zoonotic transmission event associated with a large seafood market that also traded in live wild animals, it soon became clear that efficient person-to-person transmission was also occurring.^3^

The clinical spectrum of *SARS-CoV-2* infection appears to be wide, encompassing asymptomatic infection, mild upper respiratory tract illness, and severe viral pneumonia with respiratory failure and even death, with many patients being hospitalized with pneumonia in Wuhan and elsewhere.^4^ A global pandemic has erupted due to a high proportion of asymptomatic patients coupled with a high degree of viral shedding, long incubation period, and late clinical manifestations. Prolific testing, therefore, remains one of the most effective epidemiological interventions to stop early community spread. Unfortunately, the novelty of *SARS-CoV-2* meant that no testing was immediately available making it difficult for public health officials to stay ahead of the pandemic curve.

As State Public health laboratories became backlogged, Baylor Scott and White hospital system (BSWH) collaborated with the Luminex Corporation (Austin, TX USA) to implement a *SARS-CoV-2* real-time reverse transcription polymerase chain reaction (rRT-PCR) assay using the genetic primers previously used in China to help relieve the bottleneck. BSWH laboratory was one of the first laboratories in Texas State to adopt *SARS-Co-V2* testing to assist state public health officials for tracing and tracking patients and their immediate contacts. As the pandemic continues to spread across the nation, goal of this study was to share the early clinical trends for COVID-19 in north and central regions of Texas. The aim of this study is to encourage other laboratories to consider an early start to testing during pandemics, share initial trends in this part of the world and possible impact of *SARS-CoV-2* on other seasonal respiratory viruses. This report describes the early trends of *SARS-CoV-2* infections in the central and north Texas, USA and impact of epidemiological interventions that may have led to the decrease in the incidence of seasonal respiratory virus infections

## Methods

This study was reviewed and approved by the Baylor Scott and White Health Institutional Review board (IRB # 344003)

### Study design and participants

This retrospective cohort study included two cohorts of adult inpatients from Baylor Scott & White Medical Center in Temple (Temple, TX), representing the central Texas region, and various Baylor hospitals in Dallas, TX area, representing north Texas region. For the simplicity, all hospitals within Baylor organization will be referred to as BSW hospitals (BSWH). All adult patients were prescreened according to WHO and BSWH guidelines to be eligible for *SARS-CoV-2* testing. Briefly, patients were prescreened on BSWH web portal, phone app and/or through e-visit prior to making appointment for specimen collection at one of the several designated locations. Patients were asked for travel history and any other associated symptoms such as fever, cough and shortness of breath. When clinically indicated, *SARS-CoV-2* testing was ordered by the attending physician or by other care providers.

As BSWH laboratory continues testing, this study included data from the day testing began on March 11, 2020 and until March 23, 2020. These two hospital systems within BSWH represent central and north Texas population and are limited to these regions of Texas due to community outreach. Study includes data for *SARS-CoV-2* testing from these two regions and seasonal respiratory virus testing data is limited to central Texas region.

### Data collection

Epidemiological, demographic, clinical and laboratory data were extracted from electronic medical records and laboratory information system.

### Laboratory procedures

#### *SARS-CoV-2* testing

Methods for laboratory confirmation of *SARS-CoV-2* infection were based on rRT-PCR technique approved by FDA (US Federal Drug and Food Administration) under Emergency Use Authorization (EUA)^5^. Briefly, all BSWH nasopharyngeal specimens were collected either by drive through collection sites or from inpatients using a flocked swab in Universal or Transport Media (Copan Technologies, USA). Specimens were transported at 2 - 8 °C to BSWH (Temple, TX) molecular pathology laboratory for processing and testing with less than 3 hours of transit time. BSWH (Temple, TX) molecular pathology laboratory was responsible for *SARS-CoV-2* detection in respiratory specimens by real-time reverse transcription polymerase chain reaction (rRT-PCR) methods (Luminex Corporation, Austin, TX USA).

The *SARS-CoV-2* primers were designed by manufacturer of the assay to detect RNA targets from the *SARS-CoV-2* in respiratory specimens from patients as recommended for testing by public health authority guidelines. The method employs two primers for amplifying *ORF1* gene and *N* gene from *SARS-CoV-2* virus and the assay includes extraction and internal controls built in the same cartridge. Internal sample processing controls to verify sample lysis, nucleic acid extraction, and proper system and reagent performance are built into each Luminex Extraction Cartridge. Human RNAase P was used as an internal control. Luminex Aries offers true random-access testing, however, increased demand for testing necessitated validation of a similar assay on the Luminex NxTAG platform for batched testing, this method includes additional *Envelope (E)* gene target for *SARS-CoV-2* detection. The Luminex NxTAG platform offers high throughput but on a batched processing using similar primers as Luminex Aries. Both assays had received FDA Emergency Use Authorization prior to submission of this manuscript.

### Other respiratory virus testing

BSWH utilizes respiratory virus syndromic panel, also from Luminex, for the diagnosis of upper respiratory infections. This Luminex NxTAG assay was used as previously described^6^. The assay detects *Influenza A* and *B, Respiratory Syncytial Virus (RSV), Parainfluenza 1-4, Human metapneumovirus, Rhinovirus/Enterovirus, Adenovirus, Bocavirus, Coronaviruses HKU1, NL63, 229E, OC43, Chlamydophila pneumoniae, Legionella pneumophila* and *Mycoplasma pneumoniae*. DNA/RNA is extracted using an automated extraction. This test is based on Luminex’s respiratory pathogen panel technology to amplify multiple targets within a single tube and is read on the Luminex MagPix workstation. In addition, BSWH laboratory uses standalone PCR tests for *Influenza* and *RSV* on Roche LIAT system (Roche molecular, Indianapolis, IN USA) or Luminex Aries and the tests performed on these instruments were included in the data analysis for this study. A *Chi*-square test was used to assess the association between the rate of infection for each virus between 2019 and 2020. Statistical significance was set at *p-value<0.05*.

## Results

### BSWH testing burden during initial periods of community spread

BSWH laboratory was one of the first few laboratories in the Texas State to start testing for *SARS-CoV-2* using Luminex Aries system. The assay was developed and validated for FDA approval under the Emergency Use Authorization on March 10^th^, 2020. While BSWH EUA application was under review by FDA manufacturer received FDA EUA on April 03, 2020 before this manuscript was submitted hence BSWH laboratory accepted manufacturer’s performance claims with limited internal verification. The BSWH laboratory started patient testing on March 11, 2020 and supported all the Baylor hospitals in central and north Texas regions. Data presented in figure 1 includes daily test volumes, combined from both Luminex Aries and NxTAG platforms. A total of 3,571 nasopharyngeal specimens were tested until March 23^rd^, 2020. The north Texas region contributed 1,219 while central Texas region 2,352 specimens. The typical turnaround time for specimen collection to verification of test results was less than 15 hours.

**Fig 1.**
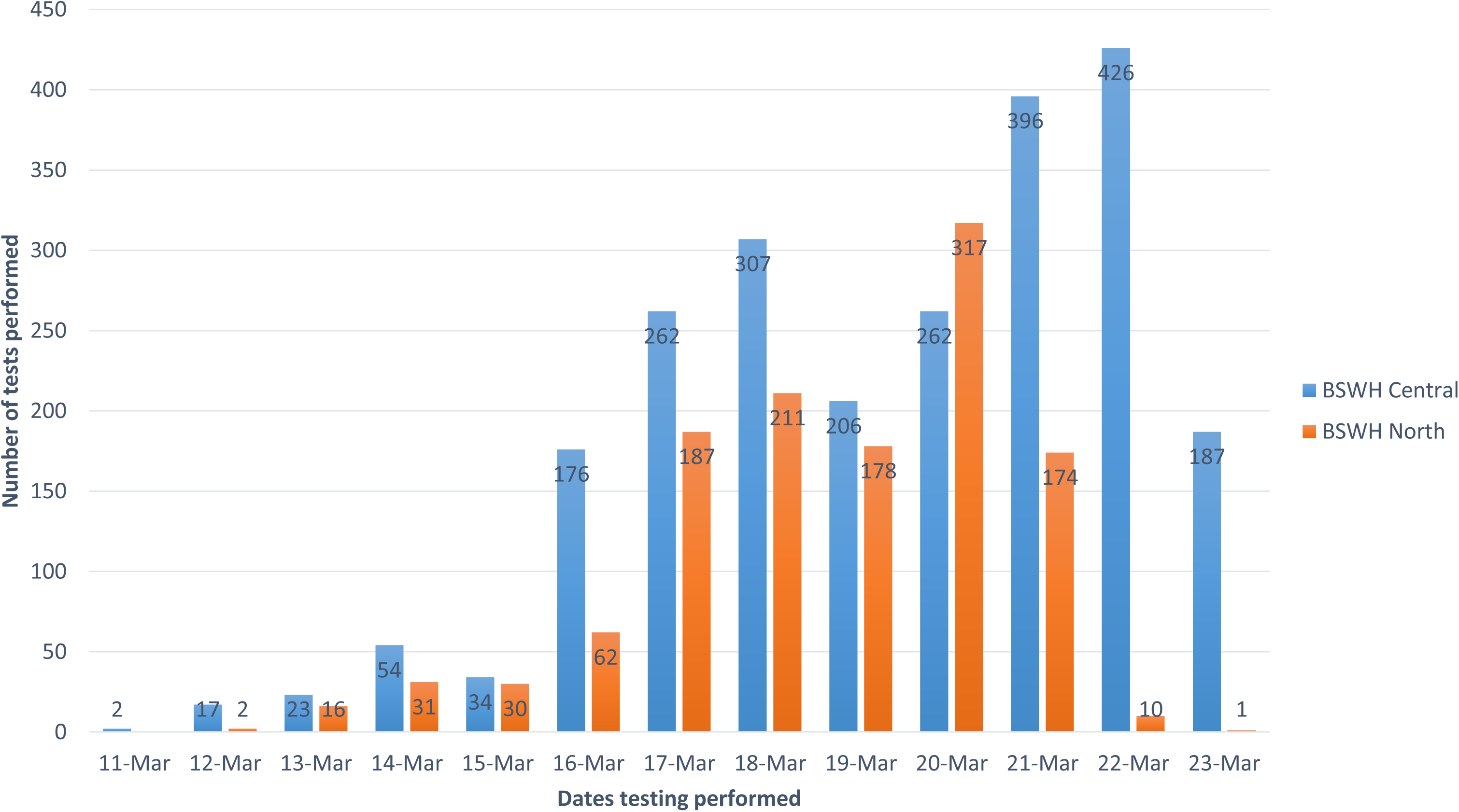
Number of SARS-CoV-2 tests performed at Baylor Scott & White Medical Center. Baylor Scott and White Memorial hospital initiated testing on March 11, 2020 following an Emergency Use Authorization submission to FDA. The data shown represents more than 3500 tests were performed between March 11, 2020 and March 23, 2020 for the two different regions of Texas State (Central and North).

**Fig 2.**
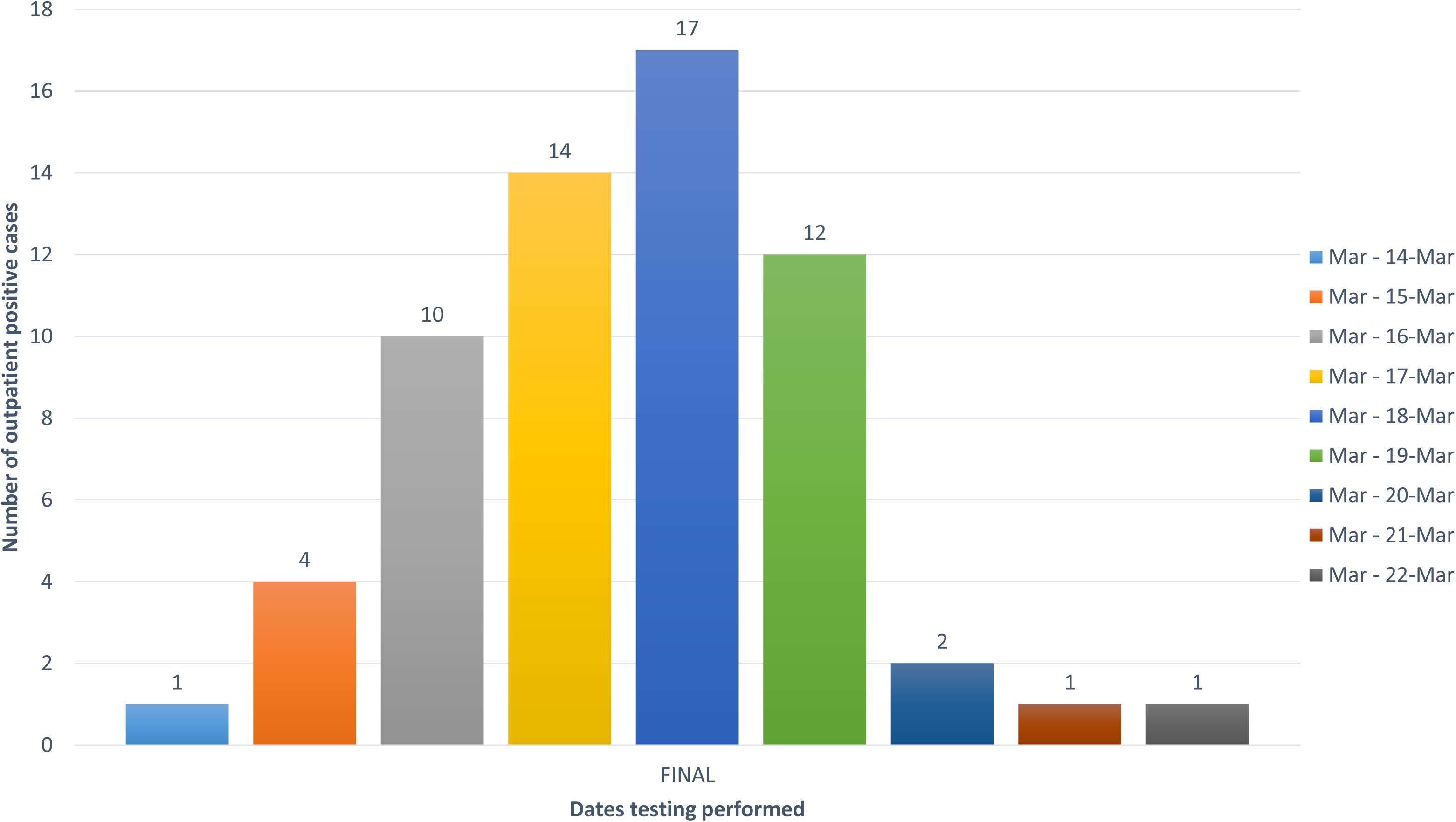
Number of positive cases in ambulatory clinics. Ambulatory patients were prescreened through a set of COVID-19 questionnaire delivered through various channels including BSWH web portal and BSWH App and/or through phone a call to be eligible for testing. Data shown are number of positive patients per day in ambulatory setting.

**Fig 3.**
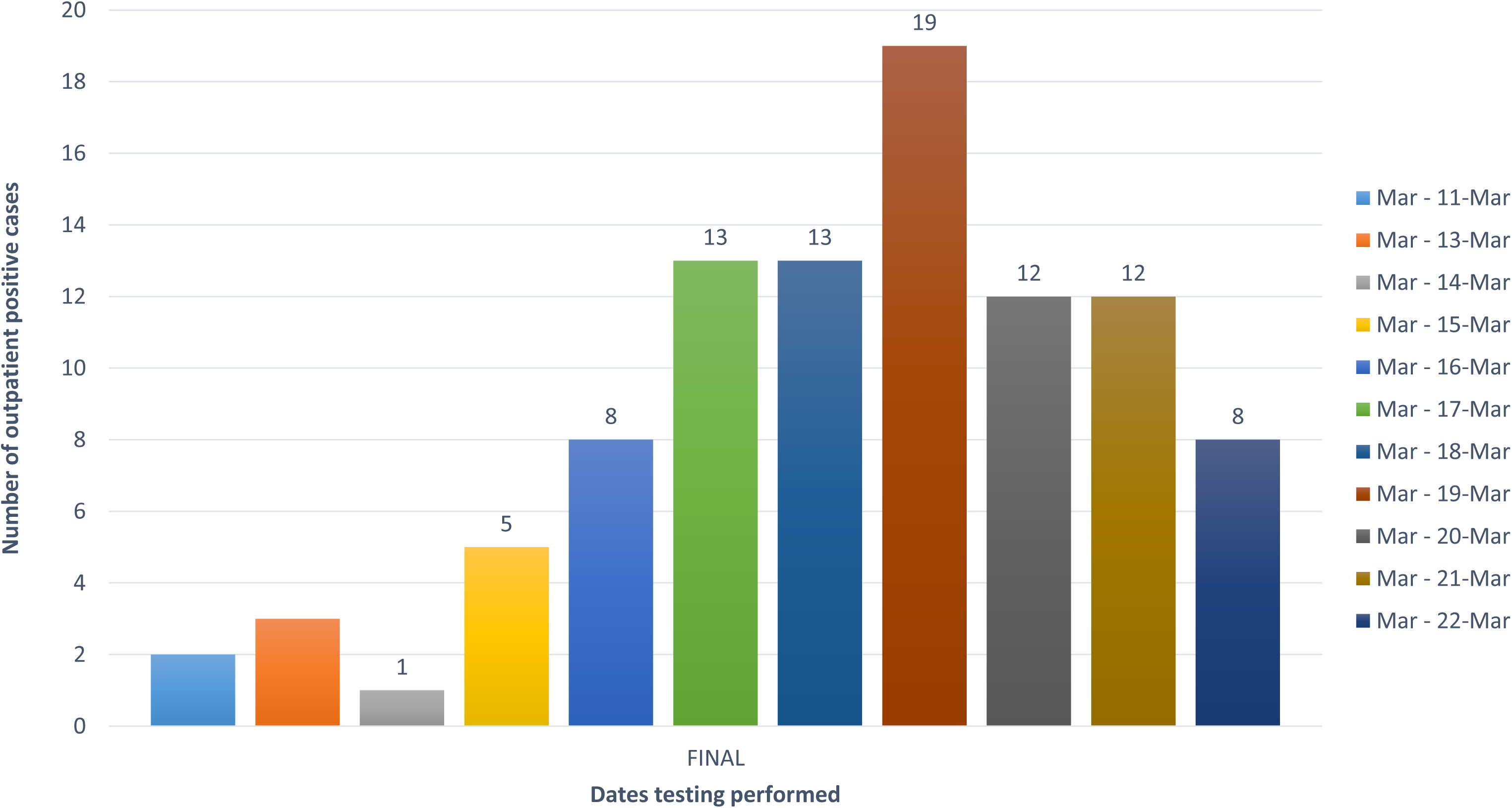
Number of positive cases in emergency department/hospitalized patients. Data shown are daily positives from March 11, 2020 to March 23, 2020. Nasopharyngeal specimens were collected from patients presented in the emergency department and/or admitted and were tested on Luminex Aries, a true random access instrument for a faster turn-around time.

### SARS-CoV-2 positive cases and age distribution

Individuals were delivered a prescreening questionnaire to be eligible for testing through BSWH web portal, phone app and/or through e-visit. The questionnaire focused on current symptoms, travel and other exposure history. Eligible patients were directed to visit one of the drive through collection sites established ad hoc for collection of nasopharyngeal specimen.

Patients with specific symptoms visiting emergency department were tested for *SARS-CoV-2* and were either discharged while waiting for test results for self-quarantine at home or admitted if clinical findings necessitated. Clinical symptoms and underlying morbidities for limited number of ED/inpatients are presented in Table 1. More than 75% of the patients presented in emergency department had fever and cough.

**Table I.** Demographic, comorbidity, symptoms, other social and past medical history of BSWH inpatient population

| Baseline characteristics (total n=14) |  |  |
| --- | --- | --- |
| Demographics |  |  |
| Gender %(n) | Male 29% (4) | Female 71% (10) |
| Age (years) | Mean 51.6 | Range 20-81 |
| BMI | Mean 32, Median 28 | Range 22-63 |
| Comorbid conditions |  |  |
| Type 2 Diabetes Mellitus |  | 28% (4) |
| Asthma %(n) |  | 36% (5) |
| COPD %(n) |  | 21% (3) |
| CHF %(n) |  | 14% (2) |
| HTN %(n) |  | 50% (7) |
| Medications |  |  |
| ACE inhibitor %(n) |  | 28% (4) |
| Angiotensin receptor blocker %(n) |  | 7% (1) |
| Social history |  |  |
| Tobacco %(n) | History of use 57% (8) | Current use 21% (3) |
| Vaping %(n) | 7% (1) |  |
| Marijuana %(n) | 7% (1) |  |
| Subjective symptoms |  |  |
| Symptom time prior (days) | Mean 4, Median 3 | Range 1-21 |
| Fever/Chills %(n) |  | 79% (11) |
| Headache %(n) |  | 36% (5) |
| Rhinorrhea %(n) |  | 28% (4) |
| Cough %(n) |  | 79% (11) |
| Otalgia/pressure %(n) |  | 21% (3) |
| Odynophagia %(n) |  | 7% (1) |
| Mild dyspnea %(n) |  | 64% (9) |
| Severe dyspnea %(n) |  | 21% (3) |
| Diarrhea %(n) |  | 7% (1) |
| Clinical characteristics |  |  |
| Abnormal Chest x-ray %(n) |  | 50% (7) |
| Peripheral pulse oximetry <94% %(n) |  | 28% (4) |
| Lymphopenia %(n) |  | 21% (3) |
| Temperature 38°C/100.4° F %(n) |  | 50% (7) |

A total of 3,571 *SARS-CoV-2* rRT-PCR tests were performed at BSWH laboratory, 1,912 specimens were received from ambulatory and/or drive-through collection sites, and 1,659 from the ED/inpatient. Sixty-two (3.2%) ambulatory patients were tested positive, as did 106 (6.3%) ED/inpatient population, noted in Figures 4 and 5, respectively.

**Fig 4.**
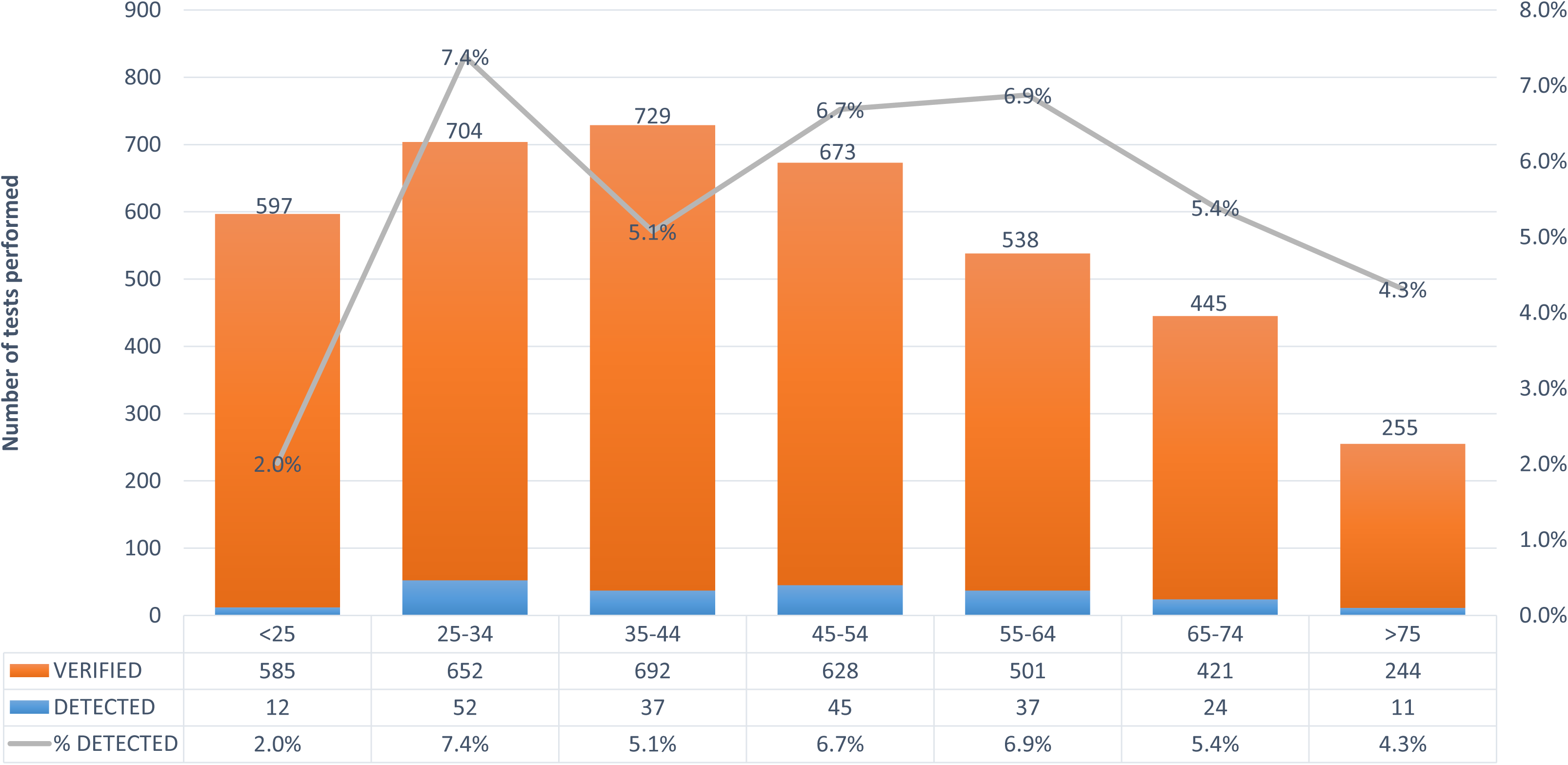
Positive test distribution in the specific group. Data shown represent positivity rate among various age groups, also shown are number of tests performed and positive results for each group.

**Fig 5.**
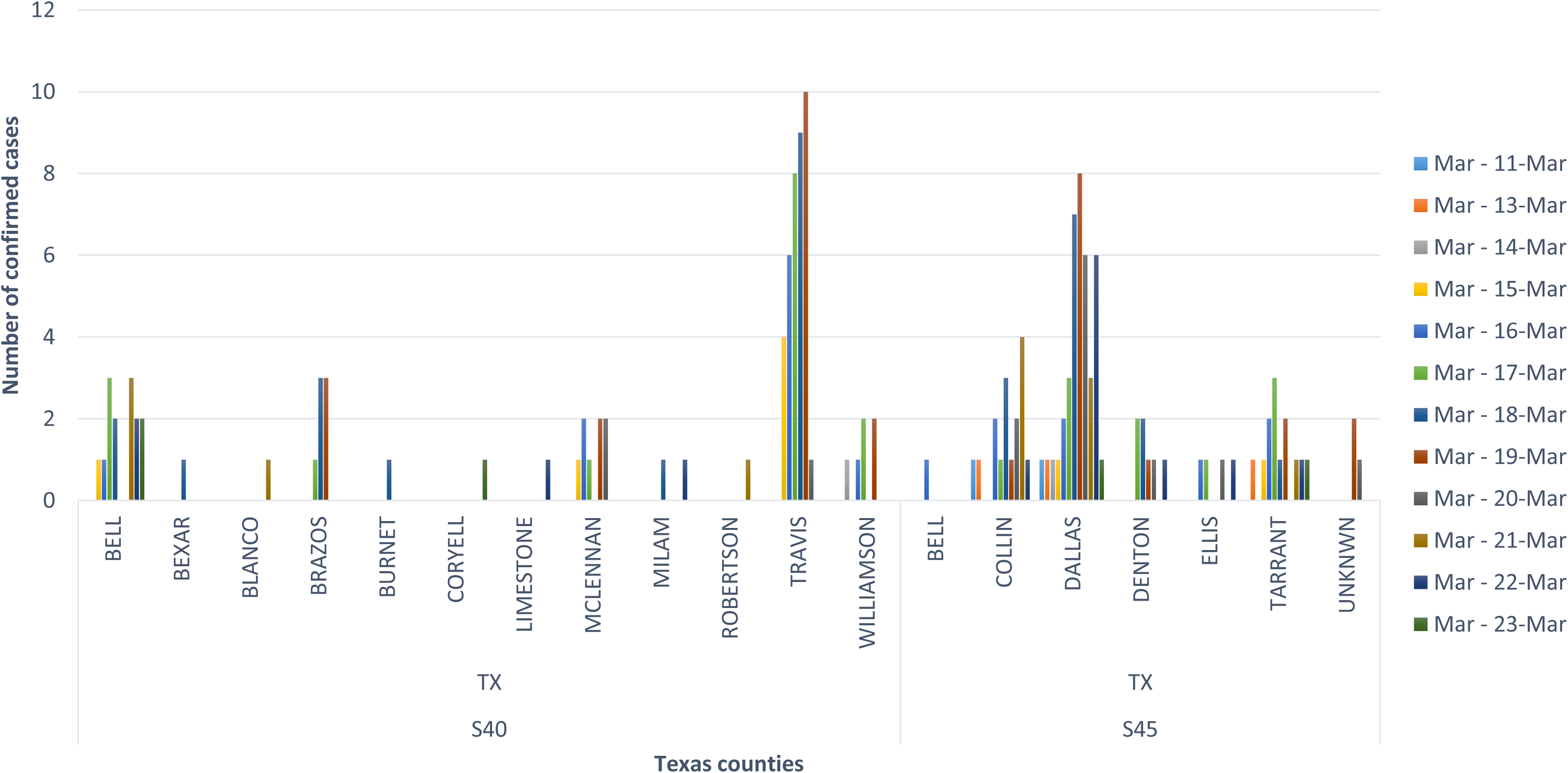
Number of confirmed cases per specific counties in north and central Texas. Epidemiological data reporting to specific public health departments. Patient demographics were extracted from the laboratory information system and segregated number of positive tests for specific counties within central and north Texas regions.

BSWH initial trends indicated a marked increase in the number of positive cases in the 25 years to 34 years age group (7.4%) followed by 6.9% in 55 years to 64 years age group. Data presented here indicated a lower incidence (2%) among the <25 years old (Figure 4).

### Number of confirmed cases per specific county in north and central Texas, USA

An appropriate epidemiological intervention requires identification of patient demographics for public health officials to track and trace positive cases. Therefore, it is prudent that *SARS-CoV-2* testing laboratories work closely with local epidemiologists for effective communication of test results. BSWH had previously built an electronic bridge with Texas Department of State Health Services for instant communication of all notifiable conditions. *SARS-CoV-2* results were added to this electronic health reporting system for an efficient communication to state epidemiologists.

Major metro areas, both in central and north Texas, witnessed an increased number of positive cases. Dallas county (north Texas) and Travis county (central Texas) had maximum number of positive cases while this manuscript was under preparation (Figure 5).

### COVID-19 impact on other circulating respiratory viruses

As local, state, and national epidemiologic countermeasures were enacted, this study observed an interesting correlation between *SARS-CoV-2* positive cases and the incidence of other seasonal circulating respiratory viruses during the same timeframe. Data extracted for BSWH from the CDC’s National Respiratory and Enteric Virus Surveillance System (NREVSS) site revealed that Influenza incidence declined to 8.7% in March 2020 compared to 25% in March 2019 (*p-<0.0001*). This declining trend over the last few weeks coincides with sharp uptick in the *SARS-CoV-2* incidence. This study also observed that *Bocavirus* and *Parainfluenza* virus infections were significantly down in March, 2020 compared to March, 2019 *(p-<0.05)*. Authors did not note a similar decline in *Adenovirus*, common cold *Coronavirus, Human Metapneumovirus, Rhinovirus and RSV* infections for March, 2020 compared to March, 2019 (Figure 6).

**Fig 6.**
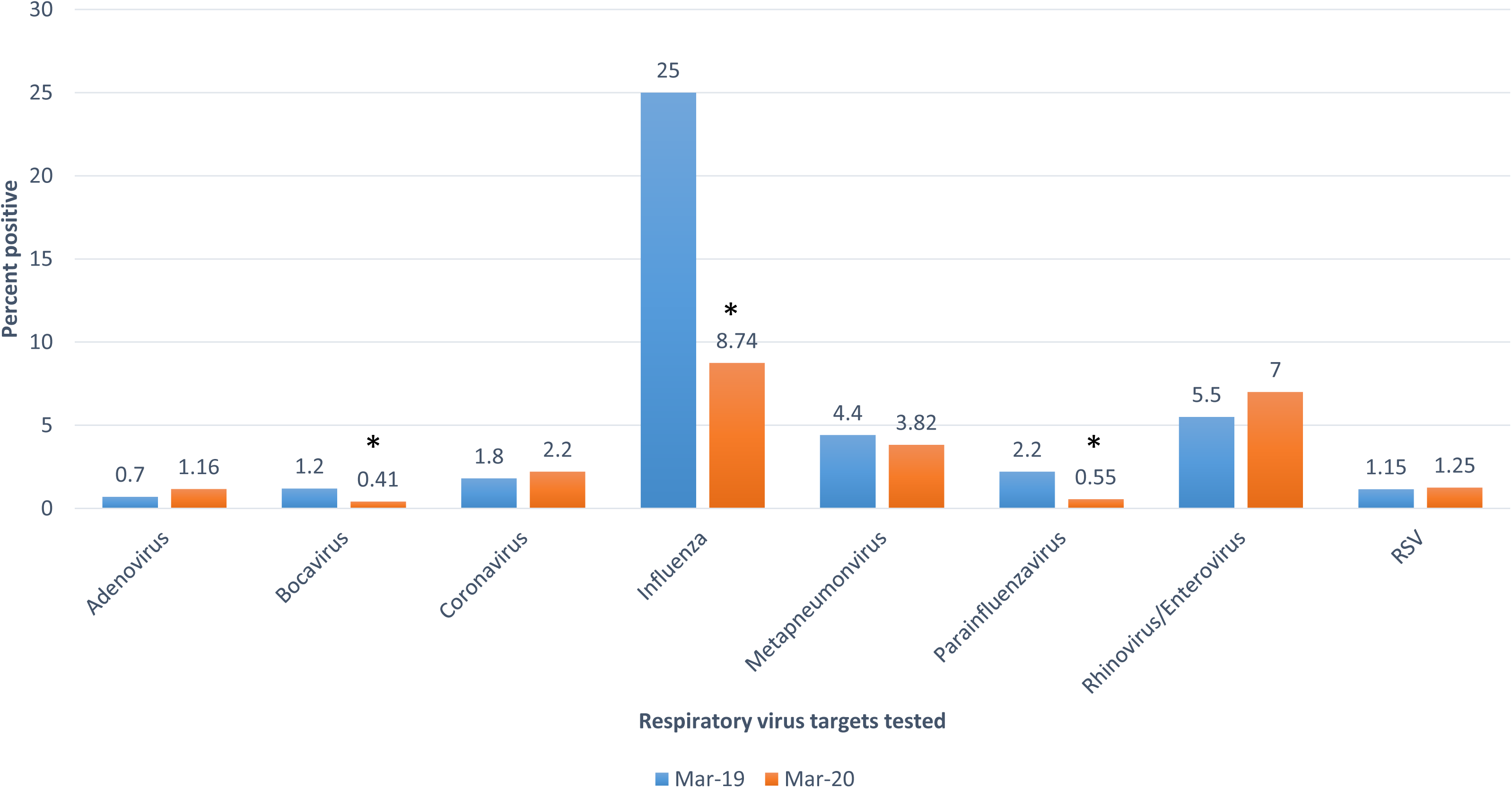
COVID-19 impact on other seasonal respiratory viruses. Decline in seasonal influenza cases. Following the enactment of epidemiological interventions by the state of Texas, large gatherings were banned and public practicing social distancing may have led to the decrease in the number of Influenza positive cases. Data shown are from March 2019 and March 2020 for percent positive test results for each virus target. A Chi-square test was used to assess the association between the rate of infection for each virus between 2019 and 2020. Statistical significance was set at *p-value<0.05*.

This study also looked at co-infections rates from *SARS-CoV-2* positive patients. We searched for 262 patient records that had concurrent testing requests for *SARS-CoV-2* and other respiratory virus infections. Contrary to several other reports from other parts of the nation, this study did not notice any co-infection cases with *SARS-CoV-2*.

## Discussion

The *SARS-CoV-2* literature is evolving at breakneck pace, but there is a paucity of literature detailing in-house testing solutions to combat the national delays in turn-around time or the shortages of testing kits available. As real-time rRT-PCR is already widely deployed in diagnostic virology laboratories, this study recommends any institution with molecular testing capabilities consider proactively reaching out to manufacturers to improve testing capabilities and turn-around time. In the race against this pandemic, real-time data empower epidemiologists and public health officials to identify, track, and contain spread as much as possible. Integrating laboratory-based reporting with epidemiologic surveillance registers will only further improve public health outcomes.

The intent of this study was not to assess the performance characters of the rRT-PCR assay for the detection of *SARS-CoV-2* infection. Authors are of the opinion that accurate determination of test performance characters will require appropriate distribution of cohorts among the general population especially in the context of virus shedding, transmission dynamics, asymptomatic carriage and specimen requirements are still being debated and investigated. *SARS-CoV-2* has exhibited great degree of plasticity in all of the above characters hence it may take additional time and understanding to determine the performance characters of the assay.

The literature data available at the time of the emergency were few for most USA healthcare systems and above all stemming from the only experience available on the outbreak from COVID 2019. The only country with published data and epidemiological or management studies was represented by the Chinese outbreak.^7^ However, the health system and the Chinese government represent a very different model from the USA reality where healthcare is regional and private for most part, which enjoys significant autonomy such as the possibilities available to try to improve and optimize diagnosis, management and partnership with public health officials. In this context, BSWH ramped up efforts in laboratory diagnosis and collegial collaboration with public health officials for effective epidemiological interventions.

Because *SARS-CoV-2* infection symptoms range from unspecific mild respiratory symptoms to acute respiratory distress^4^ and because these symptoms are very similar to those of many seasonal viruses,^8^ BSWH laboratory implemented an outpatient screening protocols hosted on BSWH web portal for appointments, phone app and e-visit sites for appropriate prescreening of individuals for targeted laboratory testing.

Real-time rRT-PCR testing for various other infections is widely deployed in most diagnostic laboratories. In the case of a public health emergency, proficient diagnostic laboratories can rely on this robust technology to establish new diagnostic tests within their routine services before pre-formulated assays become available. In addition to information on reagents, oligonucleotides and positive controls, laboratories working under quality control programs need to rely on documentation of technical qualification of the assay formulation as well as data from external clinical evaluation tests.^8, 9^ Everything listed above can be true for a laboratory-developed test, however, if commercial manufacturers design assays under FDA watch then all of the above requirements can be mitigated. The available genome sequence of *SARS-CoV-2* has enabled several diagnostic kit manufacturers to design their primer sets for real-time rRT-PCR diagnostic test builds^11^ in addition to other respiratory pathogens testing.

BSWH worked diligently with Luminex Corporation to adopt and submit an FDA emergency use authorization application of their assay for BSWH healthcare system during early phases of community spread in Texas State. This early adoption of rRT-PCR assay led to improved turnaround times of *SARS-CoV-2* test results, reduced testing burden of public health laboratory, and won praise from the local public health officials for efficient communication of test results for appropriate interventions.

To best of authors’ knowledge, this is the first report on *SARS-CoV-2* testing from this part of the world. BSWH laboratory would like to share this information with our readers and other laboratories that early adoption of testing for pandemic diseases like COVD-19 has long-term implications in management and control measures. BSWH laboratory provided test results data on both ambulatory and inpatient population, and shared patient demographics with local public health officials.

This study provided limited insights into clinical manifestations of patient who either reported to emergency department or admitted for further evaluations. Major symptoms included were fever and cough, more than 75% of the patients reported to have these symptoms. Khuwara *et al^12^* reported similar findings in Wuhan outbreak, reporting greater than 90% and 75% of the patients exhibiting fever and cough, respectively.

Interestingly, data mining did not yield any co-infections with *SARS-CoV-2* unlike Stanford Medicine data^13^. This study attributes the initial trend of not finding co-infections with *SARS-CoV-2* to limited concurrent test ordering for other respiratory viruses in an ambulatory setting. A generic notion of prohibitive cost of respiratory syndromic panels may have led to the limited ordering in outpatient testing.

The coronavirus disease 2019 has rapidly spread around the world, posing enormous health, economic, and social challenges to societies. As there are no proven drug and vaccine treatments,^14^ non-pharmaceutical measures are essential to slow the spread of the epidemic.^15^ Social distancing (e.g., cancellation of large gathering, school closures) is an essential part of public health measure for infection control.^15^ In line with this, many social events and activities have been cancelled or scaled-down in many countries including Japan,^2^ wherein there is already a high number of reported COVID-19 cases.

This study demonstrated and incidental correlation of decline in the other respiratory viruses such as *Influenza* viruses typically circulating during this time of the year. This observation may be merely coincidental, however, this study hypothesized that the general epidemiological measures such as social distancing, cancellation of large gatherings and in general population is being extra-careful in preventing *SARS-CoV-2* infection may have led to the decrease in *Influenza* cases compared to what BSWH laboratory witnessed last year around the same period of time. This study must not be intended to draw any definitive conclusion on this fact, especially when data for 2019-2020 flu vaccine effectiveness is still evolving.

## Conclusion

This study was intended to provide an initial experience of dealing with a pandemic and how laboratories are required to be part of the crisis management. This study demonstrated that proactive collaboration with assay manufacturers would enable laboratories to be prepared for emerging diseases like COVID-19. Epidemiological interventions depend on availability of accurate diagnostic tests and throughput capacity of such system during large outbreaks like *SARS-CoV-2*. It is also important to have a well-organized plan to report the test results to public health officials to initiate counter measures to control the infections. It is also imperative to build a diagnostic algorithm to include testing for other seasonal respiratory viruses, especially most common viruses like *Influenza* and *RSV*, which may require medical attention.

## Data Availability

NA

## Acknowledgment

Authors would like to thank Jeffry Hunt for help with data extraction and Courtney Shaver for help with statistics.

